# Forecasting of COVID-19 Cases and Deaths Using ARIMA Models

**DOI:** 10.1101/2020.04.17.20069237

**Authors:** Lutfi Bayyurt, Burcu Bayyurt

## Abstract

After the outbreak of severe acute respiratory syndrome (SARS-2002/2003) and middle east respiratory syndrome (MERS-2012/2014) in the world, new public health crisis, called new coronavirus disease (COVID-19), started in China in December 2019 and has spread all over countries. COVID-19 coronavirus has been global threat of the disease and infected humans rapidly. Control of the pandemi is urgently essential, and science community have continued to research treatment agents. Support therapy and intensive care units in hospitals are also efective to overcome of COVID-19. Statistic forecasting models could aid to healthcare system in preventation of COVID-19. This study aimed to compose of forecasting model that could be practical to predict the spread of COVID-19 in Italy, Spain and Turkey. For this purpose, we performed Auto Regressive Integrated Moving Average (ARIMA) model on the European Centre for Disease Prevention and Control COVID-19 data to predict the number of cases and deaths in COVID-19. According to the our results, while number of cases in Italy and Spain is expected to decrease as of July, in Turkey is expected to decline as of September. The number of deaths in Italy and Spain is expected to be the lowest in July. In Turkey, this number is expected to reach the highest in July. In addition, it is thought that if studies in which the sensitivity and validity of this method are tested with more cases, they will contribute to researchers working in this field.

## INTRODUCTION

New coronavirus disease (COVID-19) is a pandemi caused by a newly discovered *Coronavirus*. Most people from all of the world infected with the new C*oronavirus* (2019-nCoV) at the present time. *Coronaviruses* are enveloped, positive single-stranded RNA viruses that infect humans and many animals.^1^ 2019-nCoV affects different people in different ways. The common symptoms of the disease are fever, fatigue and dry cough. *Coronaviruses* have caused two major pandemics, such as SARS and MERS over the past two decades.^2,3^ Since the SARS outbreak 18 years ago, a large number of SARS-related *Coronaviruses* (SARSr CoV) have been discovered in bats. Zhou et al. (2020) reported that 2019-nCoV was identified in China (Wuhan) that caused an acute respiratory syndrome epidemic in humans.^4^

The outbreak that started on 12 December 2019 continues to spread worldwide and results in fatality. Since there is no approved treatment for COVID-19 currently prevention and preparation in healthcare services are crucial. This study include Italy and Spain that have high number of case and death from Europe also Turkey in which number of the case and death start to increase. From February 2, 2020 to March 27, 2020; means of the fatality rates of Italy, Spain and Turkey are calculated as %10, %7 and %2, respectively. Modeling and future forecast of daily number of confirmed cases and deaths can help the treatment system.^5^ Aim of present study, the statistical prediction models could be meaningfull in forecasting and controlling this global pandemic threat. For this purpose, Auto Regressive Integrated Moving Average (ARIMA) model was used to predict the confirmed cases and deaths of COVID-19.

## MATERIALS AND METHODS

The daily case and death data of COVID-19 from February 2, 2020 to March 27, 2020 were collected from the official website of European Centre for Disease Prevention and Control (https://www.ecdc.europa.eu/en/publications-data/download-todays-data-geographic-distribution-COVID-19-cases-worldwide).^6^ Excel 2019 was used to build a time-series database. SPSS 21 and Eviews 10 statistical software was used to perform statistical analysis on the case and death datasets, and the statistical significance level was set at 0.05.

ARIMA model consists of autoregressive (AR) model, moving average (MA) model and seasonal autoregressive integrated moving average (SARIMA) model.^7^ Various methods are used to estimate whether a time series is stationary. One of these methods is the Increased Dickey-Fuller (ADF) unit root test.^8^ Log transformation and differences are the preferred approaches to stationary the time series.^9^ Seasonal and nonseasonal differences were used to stationary the term trend and periodicity.^10^

The non-seasonal ARIMA model is usually denoted as ARIMA (*p, d, q*), in which *p* is the order of the autoregression (AR) component, *d* is the order of the differencing process to form a stationary times series, and *q* is the order of the moving average (MA) process.^11^ In an ARIMA model, the value of *y* at time *t* is estimated as equation (1)

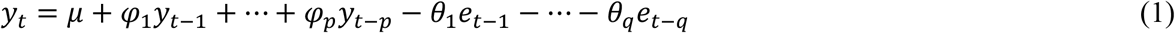

where *y*_*t*_ is the value at time *t, φ* is the AR parameter, and *θ* is the MA parameter.

Before analyzing, time series must become station-based on mean and variance. The Augmented Dickey-Fuller (ADF) is used in recognizing stationary in the mean and Ljung Box test in recognizing whether the time series is stationary based on variance or not.^8^ The stationarity levels of the series are analyzed by Dickey and Fuller (1981).^12^

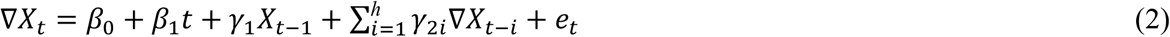

After the extended Dickey and Fuller test (ADF) expressed by equation (2) is tested by unit root test, predictions for each series are determined by ARIMA models. To determine the appropriate ARIMA (p, d, q) model, the autocorrelation and partial autocorrelation functions of each series were examined and the significance of the parameters were checked. AIC (Akaike information criterion) and BIC (Schwartz Bayes information criterion) criteria were used in the selection of the appropriate model, and the future predictions were made by choosing the model that produces the best results as a prediction model. The initial number of ARIMA model was guessed through autocorrelation function (ACF) graph and partial autocorrelation (PACF) graph. SPSS 21 and Eviews 10 statistical software were used to perform statistical analysis on the case and death datasets, and the statistical significance level was set at 0.05.

## RESULTS

Time series analysis were made for the number of cases and deaths in Italy, Spain and Turkey due to COVID-19 pandemic. When the time series graphs are examined, the trend is seen. Autocorrelation (ACF) and partial autocorrelation (PACF) graphics were used to see this more clearly and determine its stationary. In the ACF graph, it is seen that the series is not stationary since many delays exceed the confidence limits. In this case, the first order difference was applied to the series and it was ensured to become stationary ACF and PACF graphs of the first order difference of the series for Italy, Turkey and Spain are showed respectively (Fig. 1) (Fig. 2) (Fig. 3).

**Figure 1.**
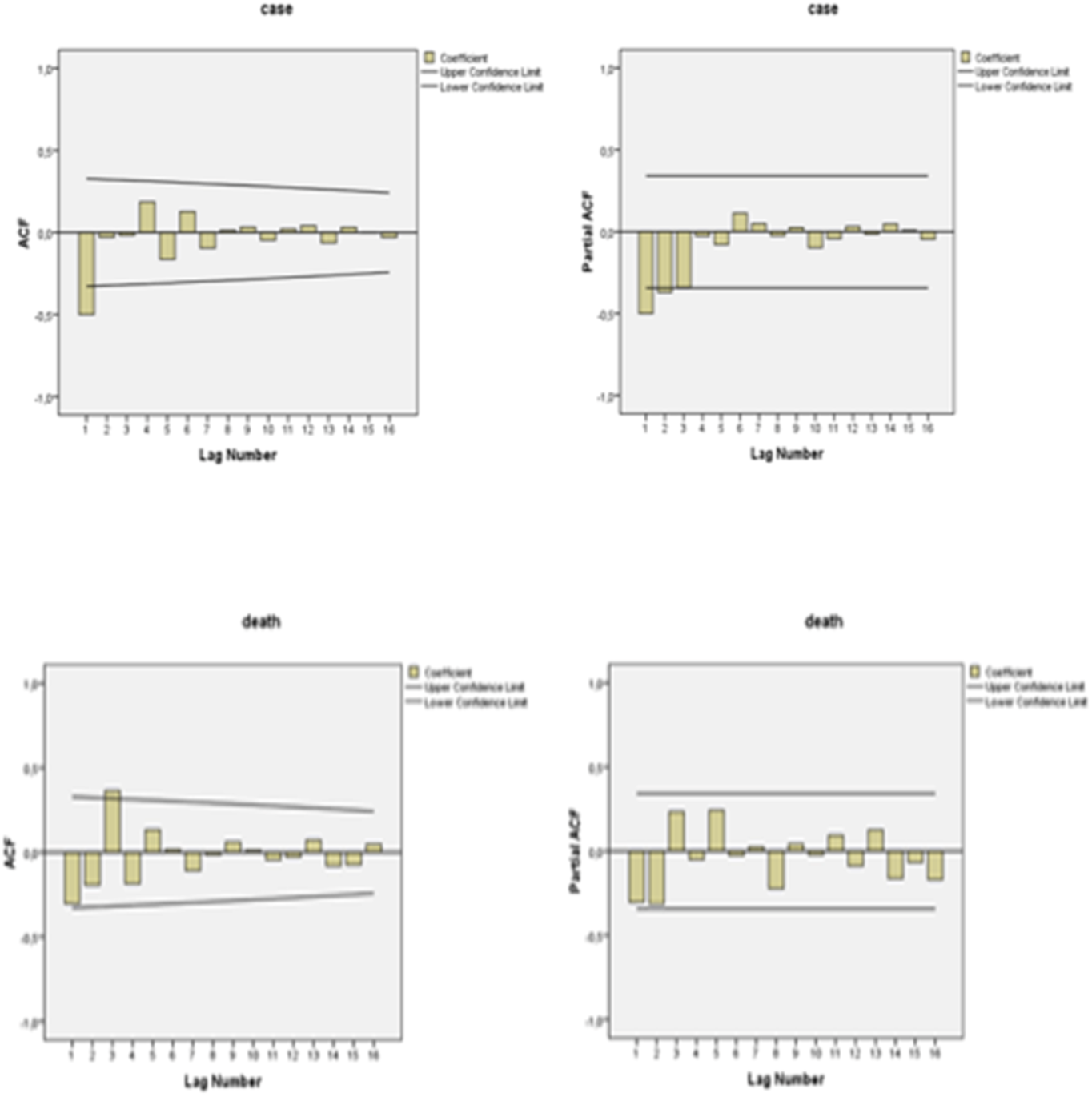
Daily confirmed cases and deaths according to first order difference ACF and PACF plots in Italy.

**Figure 2.**
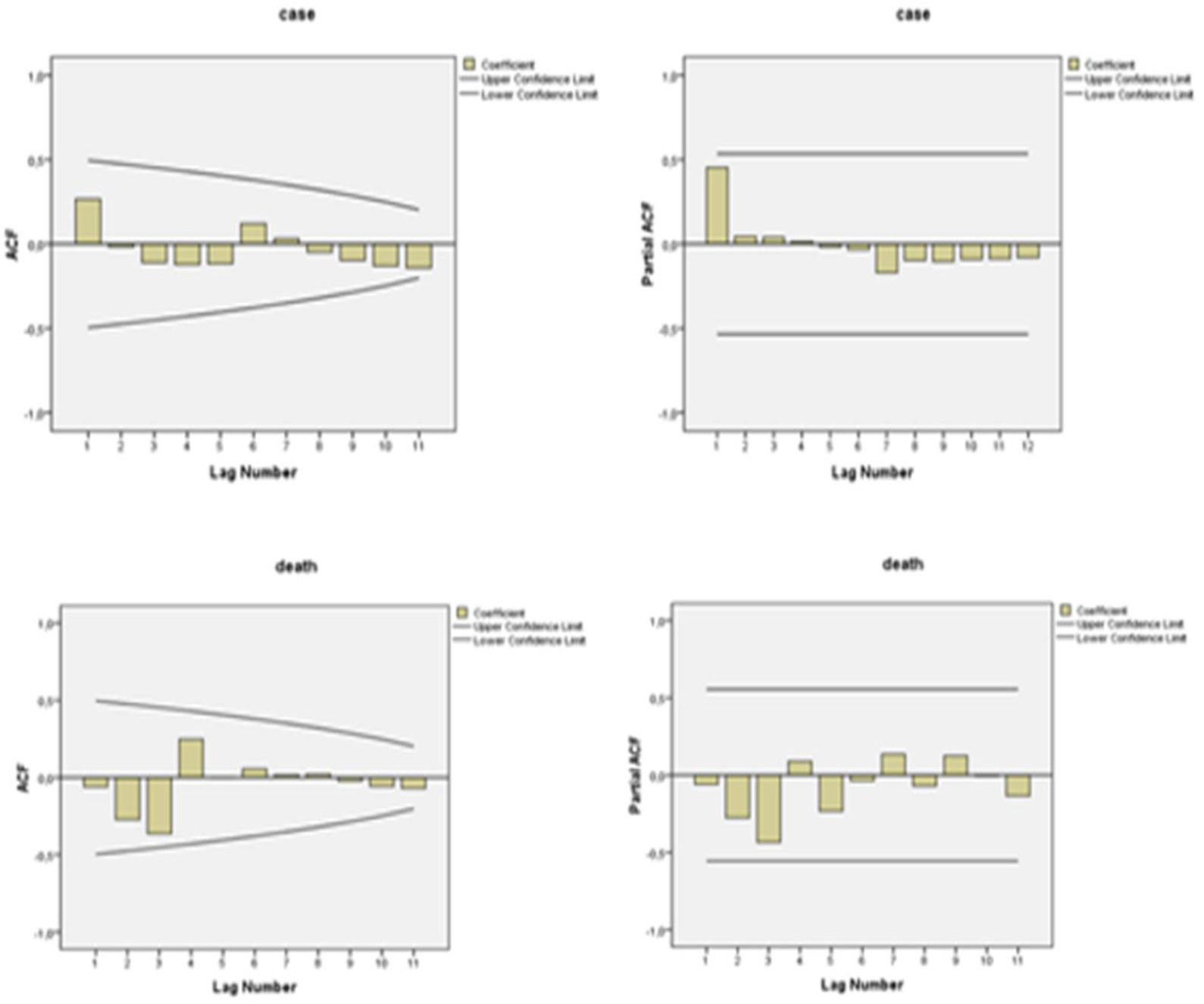
Daily confirmed cases and deaths according to first order difference ACF and PACF plots in Turkey.

**Figure 3.**
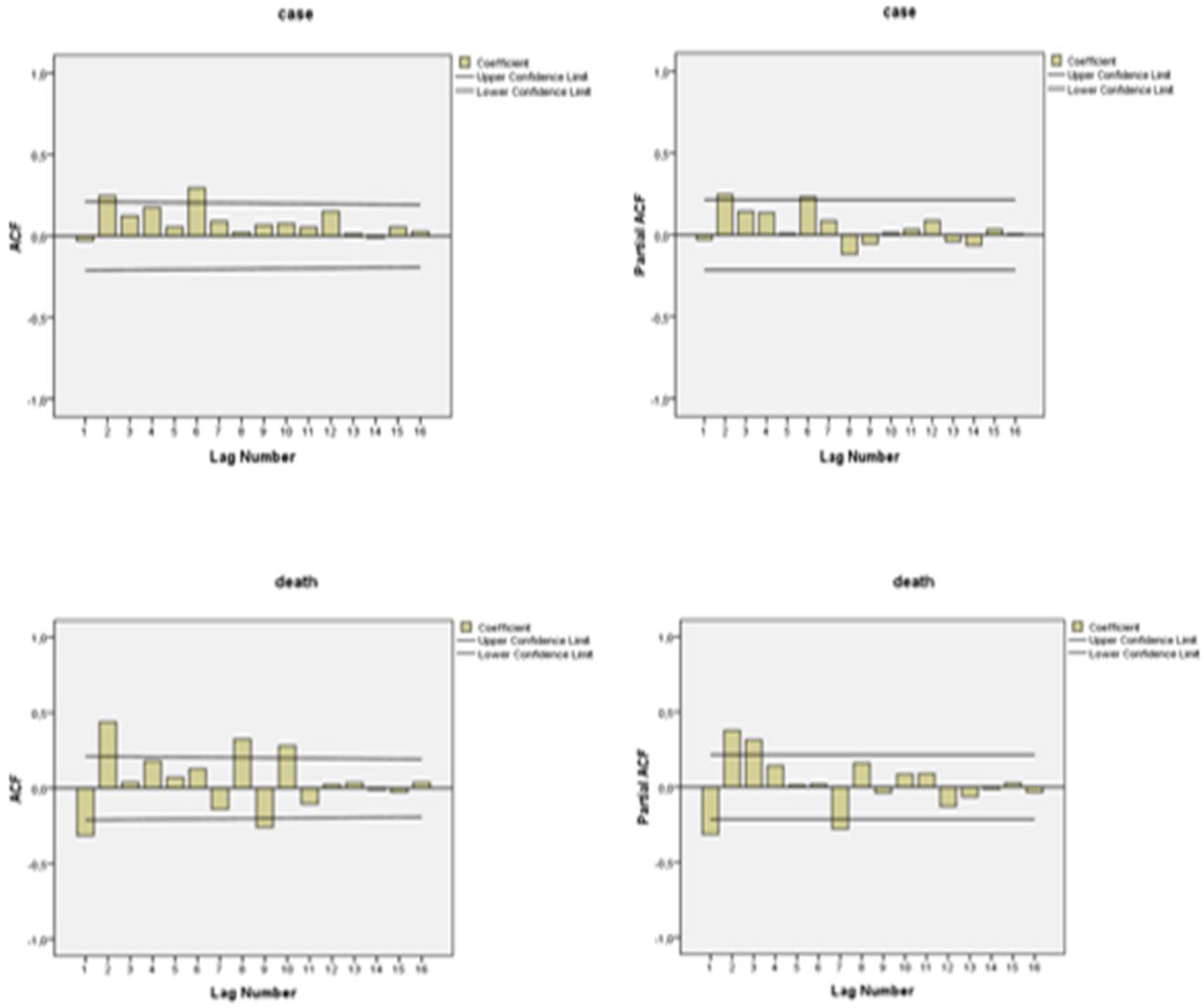
Daily confirmed cases and deaths according to first order difference ACF and PACF plots in Spain.

After the first difference procedure, the series became stationary as seen from the autocorrelation graph. Parameters of the ARIMA model were estimated by autocorrelation function (ACF) and partial autocorrelation (PACF) graph. To determine the number of case and death of COVID-19, the best ARIMA models are showed (Table I).

**Table I.**
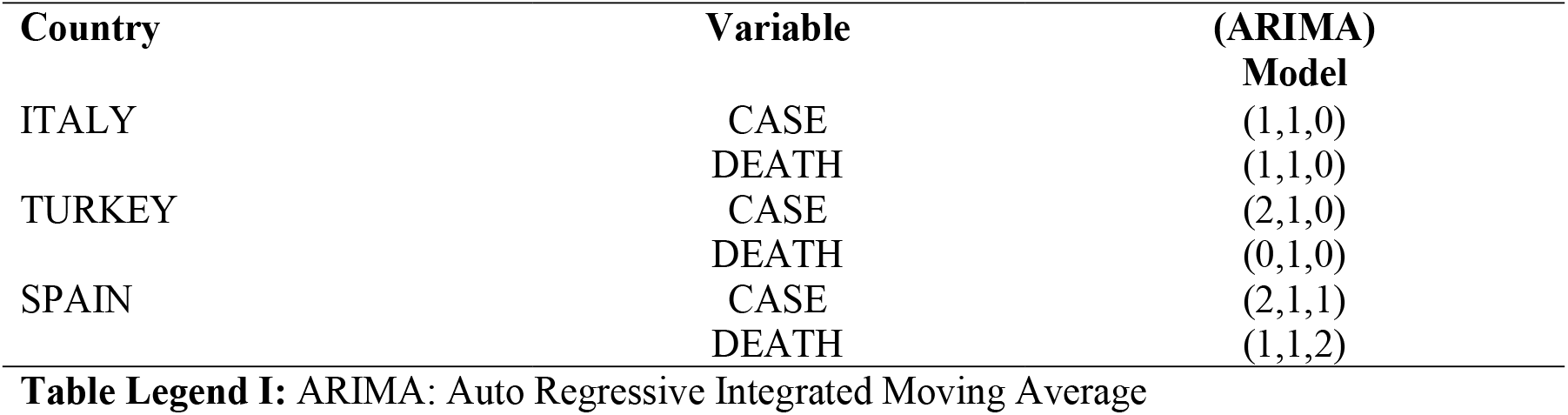
The best ARIMA models for forecasting number of daily confirmed cases and deaths according to first order difference ACF and PACF plots

| **Country** | **Variable** | **(ARIMA)**<br>**Model** |
| --- | --- | --- |
| ITALY | CASE | (1,1,0) |
|  | DEATH | (1,1,0) |
| TURKEY | CASE | (2,1,0) |
|  | DEATH | (0,1,0) |
| SPAIN | CASE | (2,1,1) |
|  | DEATH | (1,1,2) |

**Table Legend I:** ARIMA: Auto Regressive Integrated Moving Average

Time series were created using the best ARIMA models selected. Goodness of fit criteria and coefficients for the time series models are given in (Table II). According to the findings; It has been determined that the coefficients of the models used to predict the number of case and death in Italy is statistically significant (P<0.05). The constant coefficient for the number of cases and deaths was not included in the model since it was statistically insignificant (P>0.05). It was determined that the explanatory power of the estimation equation for the number of case in Italy was 76.1% and the error terms were stationary as a result of Ljung-Box statistics. It has been determined that the model can be used for foresight due to the provision of necessary assumptions. The explanatory power of the model for the number of death in Italy was calculated as 92.7%, and it was determined that the error terms were stationary as a result of Ljung-Box statistics. Models with a MAPE (Mean Absolute Percentage Error) value below 10% are very good, models with a range of 10-20% are good, models with a range of 20-50% are acceptable, and models above 50% are classified as incorrect and inaccurate. It is determined that models can be used in making future predictions due to high determination coefficients (R^2^) and MAPE value is less than 10%.

**Table II.**
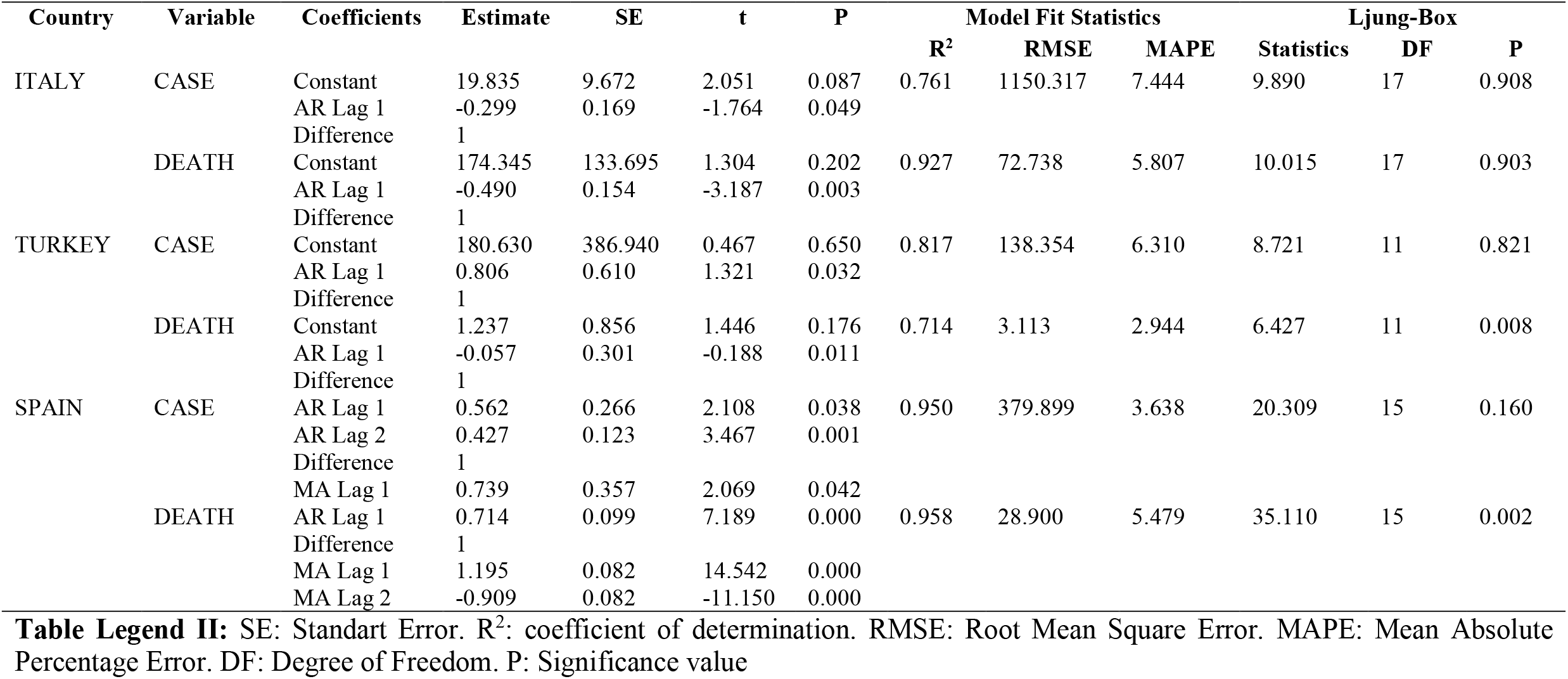
Goodness of fit criteria and coefficients for the time series model

The coefficients of the models used to predict the number of case and death in Turkey were determined to be statistically significant (P<0.05). The constant coefficient for the number of case and death was not included in the model since it was statistically insignificant (P>0.05). It was determined that the explanatory power of the estimation equation for the number of case in Turkey was 81.7% and the error terms were stationary as a result of Ljung-Box statistic. It has been determined that the model can be used for foresight due to the provision of necessary assumptions. The explanatory power of the model for the number of death in Turkey was calculated as 71.4% and it was determined that the error terms were stationary as a result of Ljung-Box statistics. it is determined that models can be used in making future predictions since MAPE value is less than 10%.

The coefficients of the models used to predict the number of case and death in Spain was statistically significant (P<0.05). It was determined that the explanatory power of the estimation equation for the number of case in Spain was 95.0%, and the error terms were stationary as a result of the Ljung-Box statistic. It has been determined that the model can be used for foresight due to the provision of necessary assumptions. The explanatory power of the model for the number of death in Spain was calculated as 95.8%, and it was determined that the error terms was stationary as a result of Ljung-Box statistics. It is determined that models can be used in making future predictions since MAPE value is less than 10%.

The confirmed and prediction graphs of the number of case and death of the countries are given in (Fig. 4-6). While number of cases in Italy and Spain is expected to decrease as of July, in Turkey is expected to decline as of September. The number of deaths in Italy and Spain is expected to be the lowest in July. In Turkey, this number is expected to reach the highest in July (Fig. 4-6).

**Figure 4.**
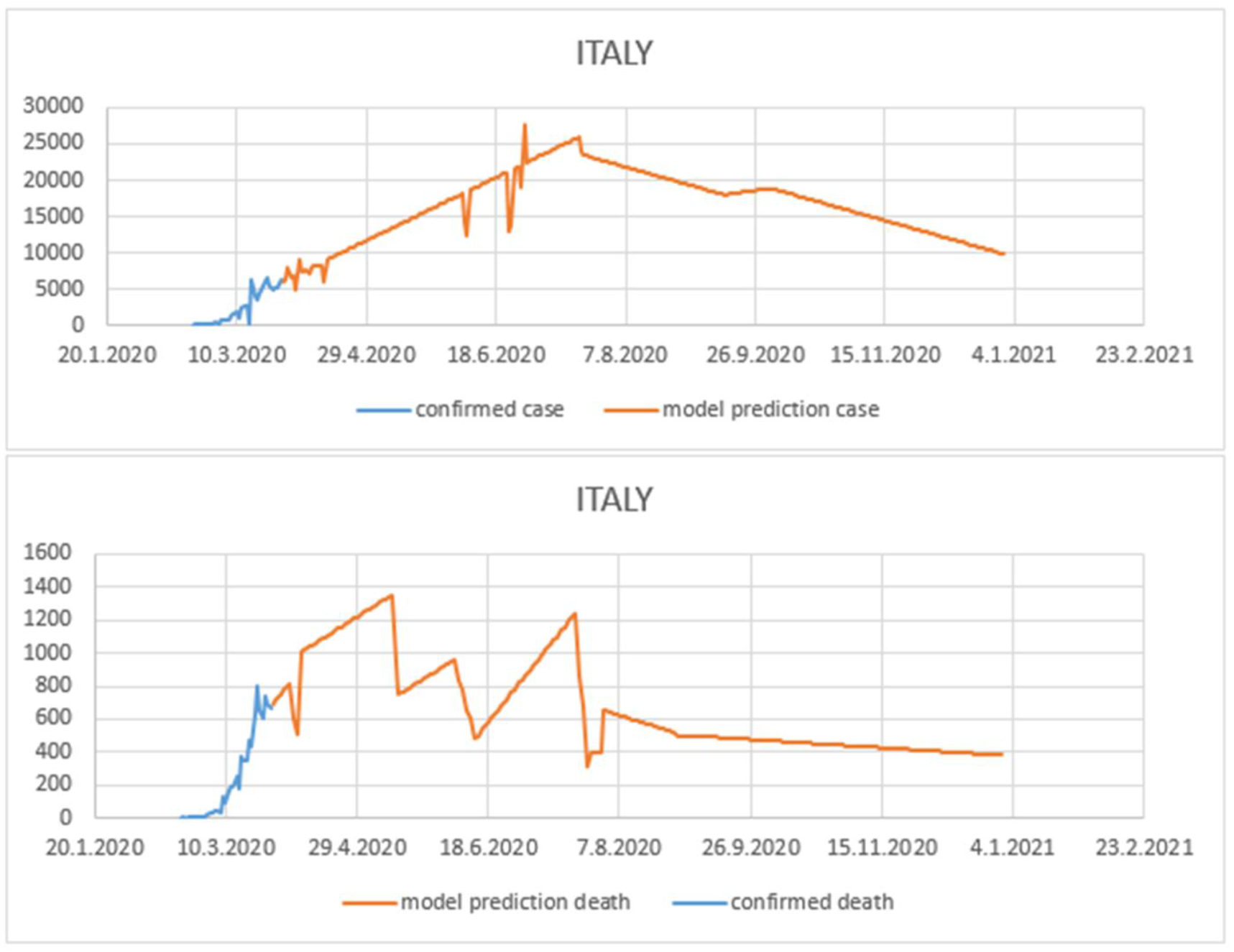
ARIMA forecast case and death graph for the COVID-19 in Italy.

**Figure 5.**
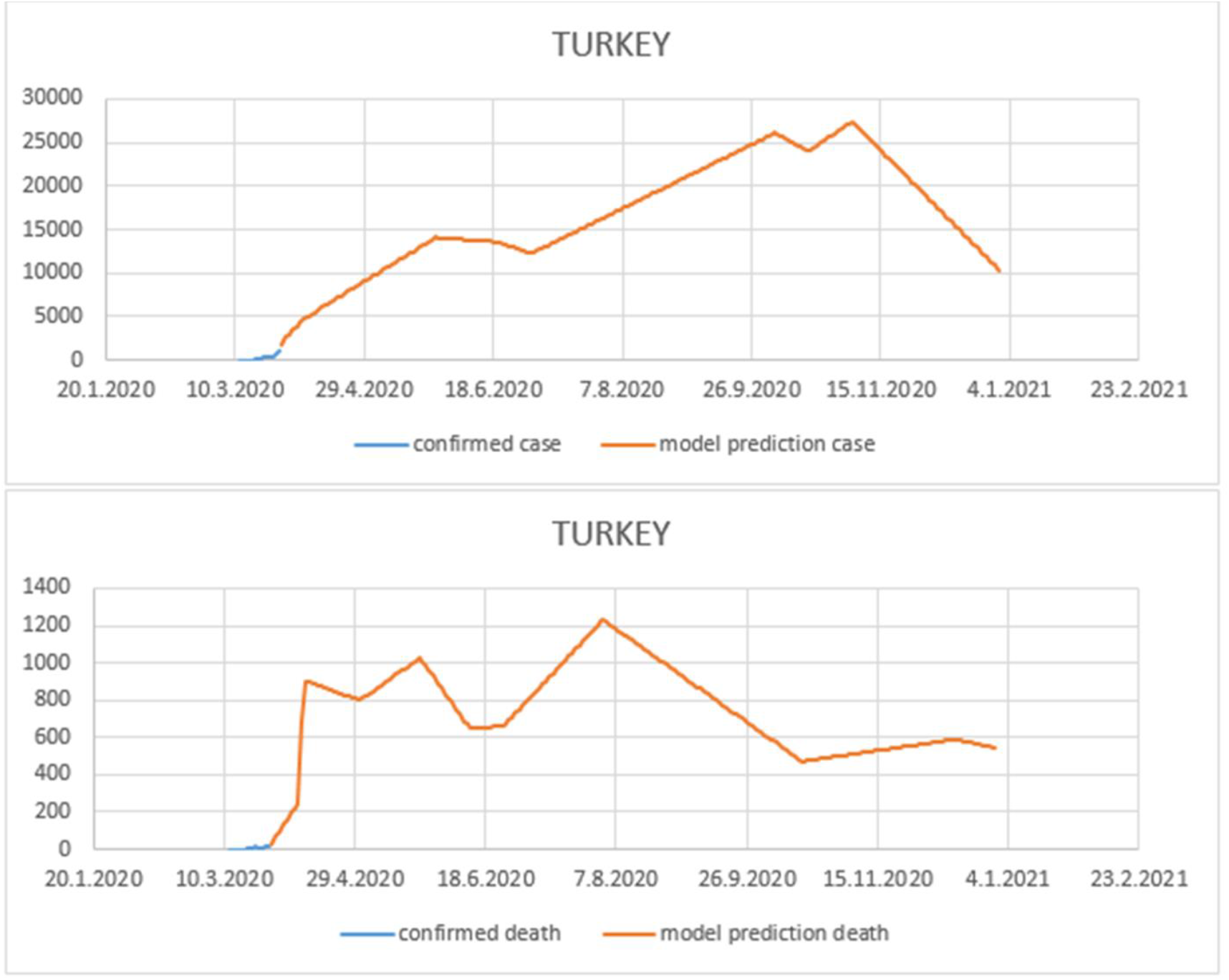
ARIMA forecast case and death graph for the COVID-19 in Turkey.

**Figure 6.**
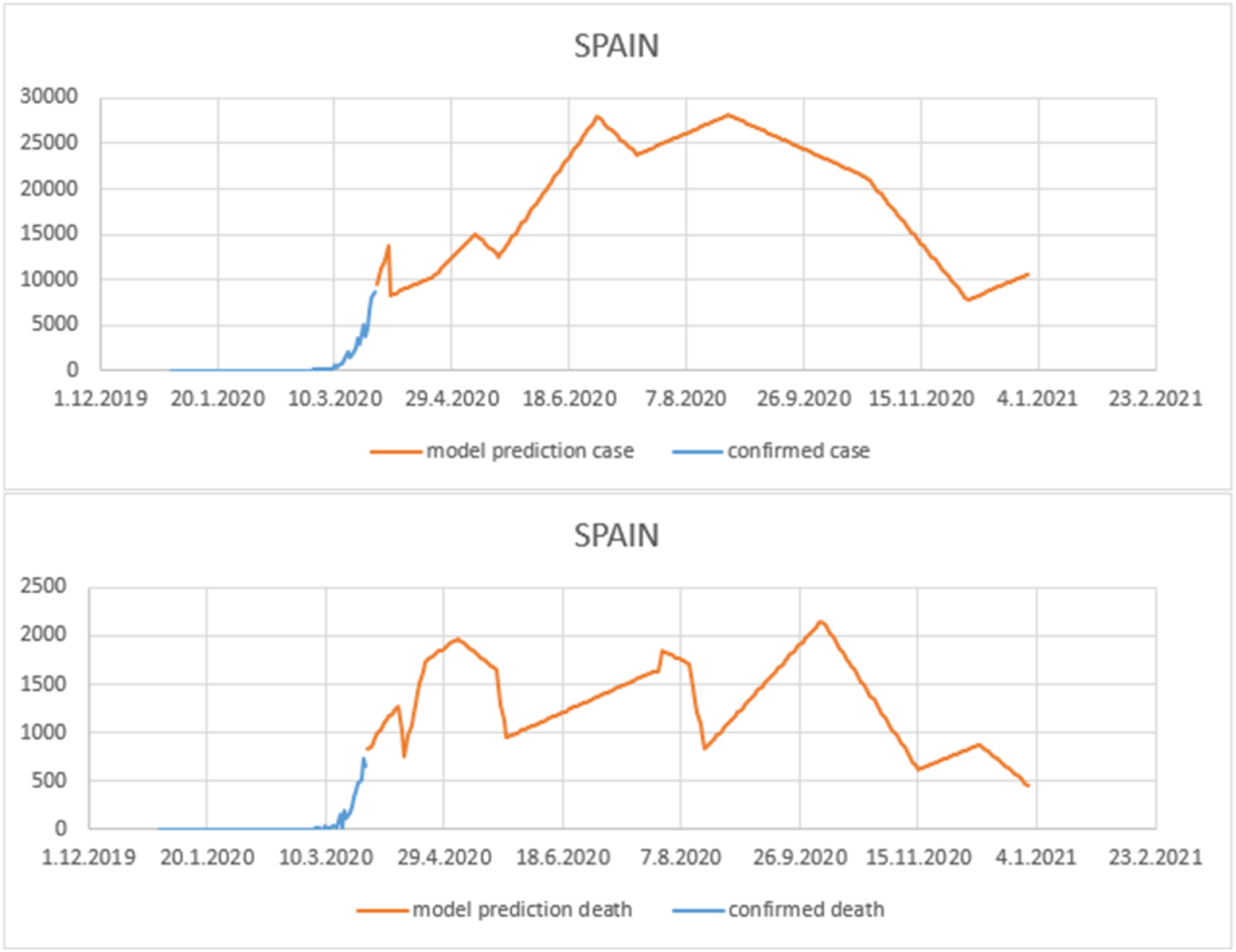
ARIMA forecast case and death graph for the COVID-19 in Spain.

## DISCUSSION

Our results are compatible with findings obtained from Benvenuto et al. (2020).^10^ In addition, (Tania et al., 2020) used ARIMA models to estimate the number of case related to the COVID-19 pandemic in Italy, China, South Korea, Iran and Thailand. It is similar to our study in estimating the number of cases in Italy.^5^ However, since the results of our study are obtained from more recent data, we think that we have obtained a more consistent forecast for the future.

In conclusion, ARIMA models have been created by considering the most appropriate AIC and BIC values for case and death numbers for each country. According to the results, while number of cases in Italy and Spain is expected to decrease as of July, in Turkey is expected to decline as of September. The number of deaths in Italy and Spain is expected to be the lowest in July. In Turkey, this number is expected to reach the highest in July. In addition, it is thought that studies in which the sensitivity and validity of these methods are tested with more cases will contribute to researchers working in this field.

## Data Availability

all data referred to in the manuscript are available

https://www.ecdc.europa.eu/en/publications-data/download-todays-data-geographic-distribution-covid-19-cases-worldwide

## REFERENCE

1. Velavan TP, Meyer CG. The COVID-19 epidemic. Trop Med Int Health, 2020;25: 278–280.

2. Drosten C, Günther S, Preiser W, Van Der Werf S., Brodt HR, Becker S, Berger. et al. Identification of a novel coronavirus in patients with severe acute respiratory syndrome. N Engl J Med, 2003;348:1967–1976.

3. Zaki AM, van Boheemen S, Bestebroer TM, Osterhaus AD, Fouchier RA. Isolation of a novel coronavirus from a man with pneumonia in Saudi Arabia. N Engl J Med, 2012;367:1814–1820.

4. Zhou P, Yang XL, Wang XG, Hu B, Zhang L, Zhang W. et al. A pneumonia outbreak associated with a new coronavirus of probable bat origin. Nature, 2020:1–4.

5. Dehesh T, Mardani-Fard HA, Dehesh P. Forecasting of COVID-19 Confirmed Cases in Different Countries with ARIMA Models. medRxiv. 2020.

6. European Centre for Disease Prevention and Control (https://www.ecdc.europa.eu/en/publications-data/download-todays-data-geographic-distribution-COVID-19-cases-worldwide)

7. Fattah J, Ezzine L, Aman Z, El Moussami H, Lachhab A. Forecasting of demand using ARIMA model. Int J Eng Bus Manag, 2018;10: 1847979018808673.

8. Cao S, Wang F, Tam W, Tse LA, Kim JH, Liu J, Lu Z. A hybrid seasonal prediction model for tuberculosis incidence in China. BMC Med Inform Decis Mak, 2013;13:56.

9. Cheung YW, Lai KS. Lag order and critical values of the augmented Dickey–Fuller test. J Bus Econ Stat, 1995;13:277–280.

10. Benvenuto D, Giovanetti M, Vassallo L, Angeletti S, Ciccozzi M. Application of the ARIMA model on the COVID-2019 epidemic dataset. Data brief, 2020;105340.

11. Shi Z, Fang, Y. Temporal relationship between outbound traffic from Wuhan and the 2019 coronavirus disease (COVID-19) incidence in China. medRxiv. 2020.

12. Dickey DA, Fuller WA. Likelihood ratio statistics for autoregressive time series with a unit root. Econometrica, 1981;1057–1072.

